# Case fatality of critically ill children treated in pediatric versus adult intensive care units in Germany: a nationwide cohort study

**DOI:** 10.64898/2026.08.14.26360448

**Authors:** Nora Bruns, Annika Wessel, Richard Biedermann, Kai Fiedler, Sarah Goretzki, Sandra Greve, Tobias Hannes, Ursula Felderhoff-Müser, Konrad Heimann, Nadine Mand, Katja Masjosthusmann, Michael Merker, Janina Soler Wenglein, Ingeborg Alijda van den Heuvel, Jens H. Westhoff, Sofia Tsaka, Victoria Lieftüchter, Christoph Härtel, Christian Dohna-Schwake, Rayan Hojeij, the Section of Pediatric Intensive Care and Emergency Medicine of the German Interdisciplinary Association of Intensive Care and Emergency Medicine (DIVI)

## Abstract

**Purpose:** Outcome consequences of critically ill children treated outside of pediatric intensive care units (PICU) are unknown. We assessed case fatality of children receiving complex intensive care treatment (CICT) by treating department in Germany and explored reasons for admission to adult intensive care units (AICU).

**Methods:** Retrospective study using the German nationwide hospital discharge dataset 2016 to 2023. Cases aged ≥ 28 days and < 18 years receiving CICT were classified as PICU, AICU, or interdisciplinary by department codes. Odds ratios (OR) for in-hospital case fatality were estimated in generalized linear mixed models with the hospital as random effect, adjusted for age, acute organ dysfunction, and chronic conditions. Excess deaths were estimated and a survey among pediatric and adult intensivists was analyzed qualitatively.

**Results:** Of 143,034 cases, 67.8 % were treated in PICUs, 14.0 % in AICUs, and 18.2 % were interdisciplinary. The crude OR for death in PICUs versus AICUs was 1.14 (95 % CI 1.03 to 1.26), reversing to 0.73 (0.63 to 0.84) after adjustment. For PICU and interdisciplinary cases combined versus AICU, the fully adjusted OR was 0.61 (0.54 to 0.70). Estimated excess deaths across the study period were 100, rising to 191 when interdisciplinary cases counted as pediatric. Capacity constraints, organizational factors, and clinical expertise were the main domains underlying AICU admissions.

**Conclusions:** Children treated outside of PICUs had higher risk-adjusted case fatality, while crude figures pointed in the opposite direction. The findings support treating critically ill children in settings with routine pediatric intensive care experience.

**Take-home message:** In a nationwide analysis of 143,034 children receiving complex intensive care treatment in Germany, risk-adjusted case fatality was higher for children treated in adult intensive care units, while the unadjusted comparison pointed in the opposite direction. Admission to adult units was driven by capacity, organizational factors, and available expertise, indicating that the place where critically ill children are treated is a modifiable determinant of outcome.

**Tweet:** Nationwide German data: children treated in adult ICUs had higher risk-adjusted case fatality than in PICUs.

## Introduction

Treatment of children in adult departments is a recurrent topic in international literature spanning emergency, trauma and intensive care. Children suffer from different diseases than adults and have age-dependent physiological needs that require specific knowledge, equipment, care logistics, and approaches to the patient [1]. Among pediatric trauma patients, mortality is higher if children are treated outside of pediatric trauma centers [2-5], which may partially be attributed to differences in decision making in end-of-life decisions [1]. Pediatric readiness describes the structural, organizational and personnel preparedness of emergency departments (ED) and other acute care infrastructures for pediatric acute care and is internationally acknowledged as key element to improve short- and long-term outcomes of children presenting to the ED [6-9]. There are numerous assessment tools available [10], but there is little evidence on the outcomes of critically ill children admitted to adult intensive care units (AICU).

For Germany, it has been shown that a relevant proportion of patients receiving complex intensive care treatment is treated outside of pediatric intensive care units (PICU) [11]. The reasons why children are treated outside of PICUs and the effects on outcomes have not been studied. However, it is conceivable that lack of pediatric readiness and expertise may exert stronger effects on outcomes of critically ill children in the ICU compared to more stable children in the non-ICU setting.

The aim of this study was to describe the distribution and characteristics of pediatric cases receiving complex intensive care treatment in pediatric, adult, and interdisciplinary settings, to assess the association between the treating department and in-hospital case fatality using the nationwide German Hospital Dataset (GHD), and to explore the reasons for pediatric admissions to AICUs in a survey among pediatric and adult intensive care specialists.

## Methods

This is a retrospective observational study using nationwide routine health care data from the GHD from 2016 – 2023. Cases were identified via age at admission and the operation and procedure (OPS) code for complex intensive care treatment (CICT) (8-98d, 8-980, 8-98f) and analyzed with respect to the treating department (PICU, AICU, or interdisciplinary (ID)).

### Data source

The GHD is a nationwide dataset that contains all discharges from public hospitals in Germany. As there are no private children’s hospitals in Germany, the dataset represents a full survey of pediatric hospitalizations in Germany. German hospitals are legally obliged to report all discharges (§21 KHEntgG). Anonymized data are made available for research at regional research data centers. We used the on-site version of the DRG statistics for the years 2016 to 2023 (EVAS 23141; DOIs 10.21242/23141.2016.00.00.1.1.0 through 10.21242/23141.2023.00.00.1.1.0). The dataset contains only the hospital stay without longitudinal follow-up.

### Case selection

Cases ≥ 28 days and < 18 years of age who received CICT and were discharged from a German hospital between 2016 and 2023 were analyzed. Coding of CICT requires specific criteria to be met and excludes cases that are admitted to the PICU for less than 24 hours and those who are merely admitted for monitoring without receiving intensive care therapies. Neonates were excluded because neonatal CICT has different coding pre-requisites and neonates are typically not admitted to AICUs.

### Determination of the treating department

The treating ICU was extracted from department codes. Cases with only pediatric department codes throughout the entire hospital stay were classified as PICU, cases with only adult department codes as AICU, and cases that received pediatric and adult department codes within the same hospital stay were classified as interdisciplinary (ID) cases. All non-pediatric department codes were considered as belonging to adult departments.

### PCCC score - Medical complexity of underlying conditions

The second version of the Pediatric Complex Chronic Conditions (PCCC) system [12] was used to determine the complexity of the case deriving from chronic conditions. It consists of 10 categories representing organ systems plus one category each for conditions originating from the neonatal period and technology dependence. We used the previously described method with the same minor modifications due to lack of information on device prescription in the GHD [11, 13].

### Pediatric Organ Dysfunction Index

Acute disease severity and treatment intensity were quantified using the Pediatric Organ Dysfunction Index (PODI), an administrative-data-based score derived from ICD-10 diagnosis and German OPS procedure codes. It was developed for administrative datasets without clinical or laboratory data and shows discrimination equal to the pSOFA to predict mortality in pediatric sepsis patients [14].

### Primary outcome

The unit of analysis was a case discharged from a German hospital between 2016 and 2023. The primary outcome was in-hospital case fatality by department.

### Understanding reasons for pediatric admissions to AICUs

To understand factors underlying pediatric AICU admissions, we developed a mini survey with three multiple-choice questions with open commentary and one open-ended question. I t was distributed among the study authors and among adult emergency and intensive care specialists to capture both perspectives.

### Missing data

Age, main diagnosis codes, and department codes were complete. Missing procedure codes cannot be distinguished from procedures that were not performed. However, CICT and organ replacement therapies are highly relevant for reimbursement, making missing codes unlikely. The same applies to secondary diagnoses, which we assumed to be coded as they increase reimbursement.

### Statistical Analysis

Frequencies were summarized as counts and percent, whereas continuous variables were presented as median and interquartile range (IQR) if skewed and mean ± standard deviation (SD) if normally distributed. Descriptive analyses were conducted overall and with the treating department and age categories (0 – 5 years, 6 – 11 years, 12 – 17 years) as grouping variables. The distribution of hospitals by the combination of departments they admitted CICT cases to, and the corresponding case volumes, was also assessed descriptively.

Crude and adjusted ORs with 95 % confidence intervals (CI) were calculated for in-hospital death with the treating ICU as exposure variable. Because it was unclear how ID cases should be classified — given that pediatric expertise was involved in their care and might also have been applied during the ICU stay — all analyses were conducted twice: comparing only AICU versus PICU cases, and AICU versus PICU + ID cases, with ID cases classified as pediatric in the latter comparison.

Multivariable hierarchical logistic regression using a generalized linear mixed model (GLMM) was performed with the hospital (identified via the institutional identifier) as a random effect. We corrected for the treating institution, because cases within a hospital are not independent with respect to central resources such as surgical expertise and equipment, even though they may be treated in different ICUs. The minimally sufficient adjustment set (MSAS) for the fixed effects was derived from a causal diagram [15, 16] based on the theory of directed acyclic graphs (DAG) [17] and included age in years (continuous), acute organ dysfunction/treatment intensity (continuous, operationalized via the PODI ) and chronic conditions (operationalized via PCCC categories) (Supplementary figure 1).

**Figure 1.**
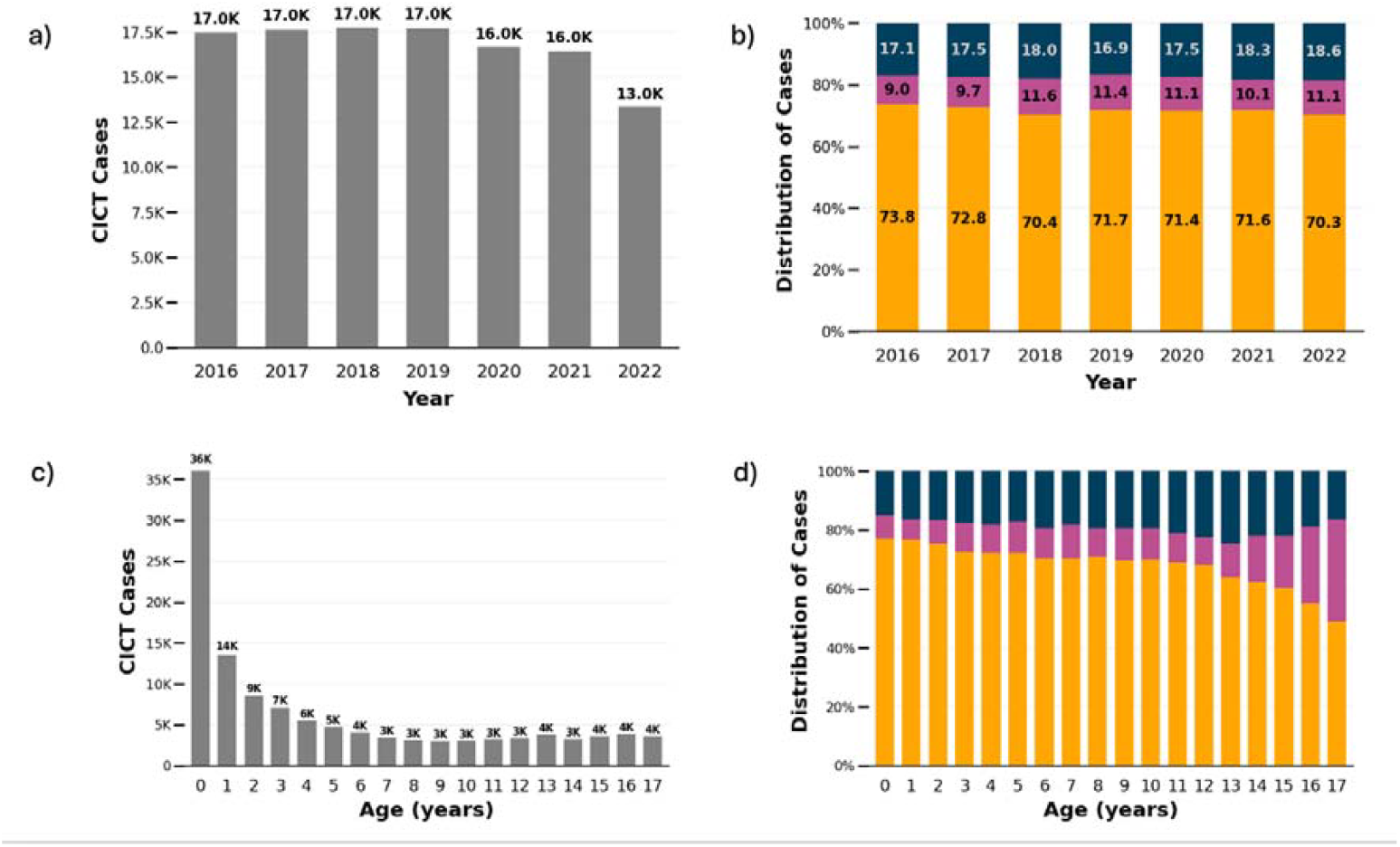
Pediatric cases in Germany receiving complex intensive care treatment (CICT) between 2016 and 2023 a)CICT cases by year b)Relative distribution of CICT cases between PICU (yellow), AICU (pink), and ID cases (blue) by year c)CICT cases by age d)Relative distribution of CICT cases between cases treated in PICU (yellow), AICU (pink), and ID cases (blue) AICU = adult intensive care unit, PICU = pediatric intensive care unit, ID = interdisciplinary cases

Excess case fatality associated with treatment outside a PICU was estimated from two counterfactual scenarios: one in which all patients were assumed to have been treated in a PICU and another with treatment in an AICU. The expected case fatality under each scenario was estimated by fitting the same GLMM as above but specified with a Poisson distribution and log link to directly estimate risks. The fitted model was applied to each counterfactual population to predict the corresponding case fatality and excess deaths were estimated from the difference of the predicted risks.

The mini survey was assessed qualitatively by categorizing insights on organizational aspects and implications of pediatric AICU admissions for the affected personnel into categories.

### Software

SAS Version 9.4 (SAS Institute, Cary, USA) was used to analyze data at the regional research data center. The DAG was created using dagitty [18] and all other visualizations were created using Python 3.13 (Python Software Foundation, Beaverton, USA) in a Jupyter Lab environment (Version 4.3.4; Project Jupyter) [19].

### Ethics approval

No ethics approval was required according to local legislation because we used exclusively anonymized secondary health care data.

## Results

The original dataset contained 15,251,642 cases, of which 143,034 met the inclusion criteria. Overall, 67.8 % of cases were treated in PICUs, 14.0 % in AICUs, and 18.2 % were interdisciplinary (Table 1). The total number of cases per year decreased from 20,306 in 2016 to 14,327 in 2023 (Figure 1a). During the same period, PICU cases decreased from 14,003 (69.0 %) to 9,097 (63.5 %) while AICU cases relatively increased from 13.3 % to 15.8 % but absolutely decreased (2,695 to 2,266) (Figure 1b).

**Table 1.** Case characteristics by treating department (pediatric hospital discharges in Germany, 2016 – 2023)

|  | Total | Pediatric | Adult | Interdisciplinary |
| --- | --- | --- | --- | --- |
| Total, *n* (row %) | 143,034 | 96,933 (67.8 %) | 20,069 (14.0 %) | 26,032 (18.2 %) |
| Sex |  |  |  |  |
| Female, *n* (%) | 62,854 (43.9 %) | 42,910 (44.3 %) | 8,694 (43.3 %) | 11,250 (43.2 %) |
| Male, *n* (%) | 80,180 (56.1 %) | 54,023 (55.7 %) | 11,375 (56.7 %) | 14,782 (56.8 %) |
| Age categories |  |  |  |  |
| 0 – 5 years, *n* (%) | 83,732 (58.5 %) | 62,926 (64.9 %) | 6,981 (34.8 %) | 13,825 (53.1 %) |
| 6 – 11 years, *n* (%) | 22,487 (15.7 %) | 15,645 (16.1 %) | 2,395 (11.9 %) | 4,447 (17.1 %) |
| 12 – 17 years, *n* (%) | 36,815 (25.7 %) | 18,362 (18.9 %) | 10,693 (53.3 %) | 7,760 (29.8 %) |
| Mechanical ventilation, *n* (%) | 48,098 (33.6 %) | 35,025 (36.1 %) | 5,582 (27.8 %) | 7,491 (28.8 %) |
| Duration of mechanical ventilation (hours), median (IQR) | 84 (35 – 192) | 88 (38 – 192) | 72 (27 – 192) | 80 (30 – 189) |
| Surgery, *n* (%) | 69,308 (48.5 %) | 44,765 (46.2 %) | 10,866 (54.1 %) | 18,095 (69.5 %) |
| ECMO, *n* (%) | 1,300 (0.91 %) | 828 (0.85 %) | 208 (1.04 %) | 264 (1.01 %) |
| Dialysis, *n* (%) | 1,613 (1.1 %) | 1,222 (1.3 %) | 136 (0.7 %) | 255 (1.0 %) |
| PODI sum, median (IQR) | 0 (0 – 1) | 0 (0 – 1) | 0 (0 – 1) | 0 (0 – 1) |
| PODI = 0, *n* (%) | 87,974 (61.5 %) | 56,799 (58.6 %) | 13,787 (68.7 %) | 17,388 (66.8 %) |
| PODI = 1, *n* (%) | 45,978 (32.1 %) | 33,878 (35.0 %) | 5,160 (25.7 %) | 6,940 (26.7 %) |
| PODI ≥ 2, *n* (%) | 9,082 (6.4 %) | 6,256 (6.5 %) | 1,122 (5.6 %) | 1,704 (6.6 %) |
| PCCC score, median (IQR) | 1 (1 – 3) | 1 (1 – 3) | 1 (0 – 2) | 1 (1 – 3) |
| PCCC = 0, *n* (%) | 31,730 (22.2 %) | 19,893 (20.5 %) | 5,609 (28.0 %) | 6,228 (23.9 %) |
| PCCC = 1, *n* (%) | 43,406 (30.4 %) | 29,113 (30.0 %) | 4,361 (35.4 %) | 7,651 (29.4 %) |
| PCCC ≥ 2, *n* (%) | 67,898 (47.5 %) | 47,972 (49.4 %) | 5,351 (43.3 %) | 12,153 (46.7 %) |
| **Outcomes** |  |  |  |  |
| Case fatality overall, *n* (%) | 3,465 (2.42 %) | 2,544 (2.62 %) | 465 (2.32 %) | 456 (1.75 %) |
| 0 – 5 years, *n* (%) | 2,035 (2.43 %) | 1,662 (2.64 %) | 138 (1.98 %) | 235 (1.70 %) |
| 6 – 11 years, *n* (%) | 539 (2.40 %) | 420 (2.68 %) | 58 (2.42 %) | 61 (1.37 %) |
| 12 – 17 years, *n* (%) | 891 (2.42 %) | 462 (2.52 %) | 269 (2.52 %) | 160 (2.06 %) |
| Length of stay (days), median (IQR) | 9 (4 – 16) | 9 (4 – 16) | 7 (3 – 13) | 10 (6 – 17) |

ECMO = extracorporeal membrane oxygenation, IQR = interquartile range, PCCC = pediatric complex chronic conditions system, PODI = pediatric organ dysfunction index

Case numbers by age were U-shaped, with 39,607 cases among infants < 1 year, three to four thousand in school children, and a slight increase from 10 years onwards (Figure 1c). The distribution of cases between departments showed a clear association with age, with older children and adolescents being treated outside of PICUs in a relevant proportion of cases (Figure 1d). Most cases (77.3 %) were treated at the 616 hospitals that admitted CICT cases to PICU, AICU, and interdisciplinary settings over the study period, while hospitals with only one department type each contributed comparatively few cases (Supplementary Table 1).

Gender distribution did not differ between treating departments, while the hospital length of stay was shortest in AICUs (Table 1) and longest in ID cases. The case fatality was generally low with 2.4 % and was highest in PICUs (2.6 %), followed by AICUs (2.3 %) and lowest in ID cases (1.8 %). The majority of deaths occurred in infants and children aged 0 to 5 years (Table 1).

Organ dysfunction codes were rarely used, with one positive category in about one third of cases and two or more positive categories in only 6.4 %. Chronic conditions were common: 22.2 % had none (PCCC = 0) and 47.5 % had two or more categories positive (Supplementary figure 2). PCCC category profiles differed markedly between unit types and age groups. The most frequently positive categories in PICU and ID cases were respiratory, cardiologic, technology dependence, and neurological conditions. In contrast, the most frequent chronic conditions in AICUs were cardiologic and were most common in young children (Supplementary figure 2).

**Figure 2.**
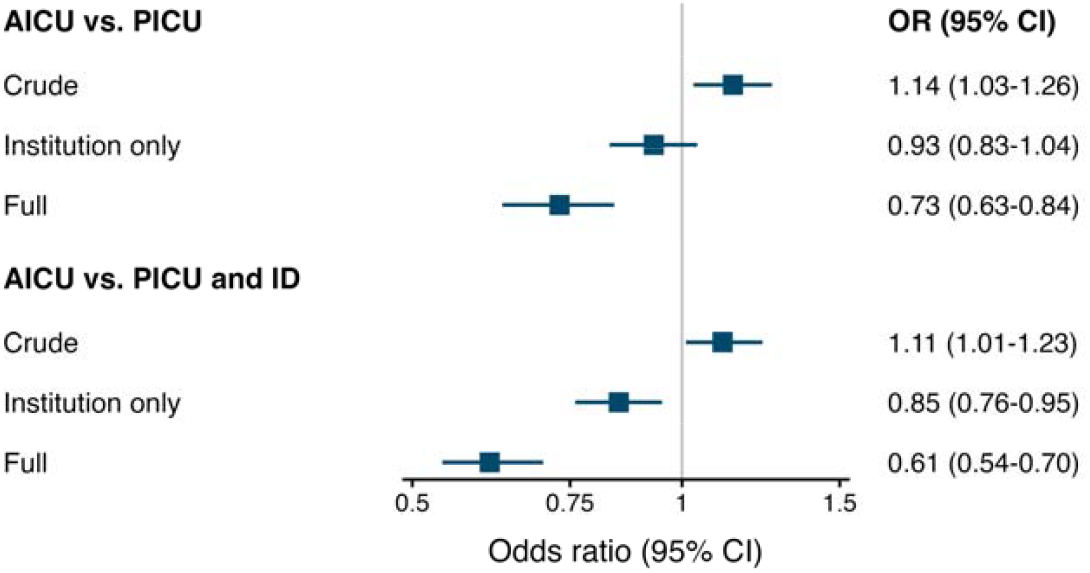
Odds ratios for case fatality by department. AICU = adult intensive care unit, PICU = pediatric intensive care unit, ID = interdisciplinary cases, PODI = pediatric organ dysfunction index, PCCC = pediatric complex chronic conditions system, OR = odds ratio, CI = confidence interval

The crude odds ratio (OR) for death in PICUs versus AICUs was 1.14 (1.03 to 1.26), changing to 0.93 (0.83 – 1.04) after adjustment for the institution as random effect (Figure 2). Additional adjustment for acute severity, chronic conditions, and age yielded an OR of 0.73 (0.63 – 0.84). For PICU + ID versus AICU cases, the corresponding ORs were 1.11 (1.01 – 1.23), 0.85 (0.76 – 0.95), and 0.61 (0.54 – 0.70).

The counterfactual analysis yielded 100 excess deaths for AICU treatment, rising to 191 when ID cases were counted as pediatric (Table 2).

**Table 2.** Excess deaths.

|  | Cases AICU | Predicted deaths (PICU scenario) | Predicted deaths (AICU scenario) | Excess deaths* | Estimated avoidable deaths |
| --- | --- | --- | --- | --- | --- |
| AICU vs. PICU | 20,069 | 2.47 % | 2.97 % | 0.50 percentage points | 100 |
| AICU versus PICU + ID | 20,069 | 2.34 % | 3.29 % | 0.95 percentage points | 191 |

\*difference between scenarios

The survey showed that reasons for pediatric admissions to AICUs are heterogeneous in origin, with capacity constraints, organizational factors, and clinical expertise as the main domains (Figure 3).

**Figure 3.**
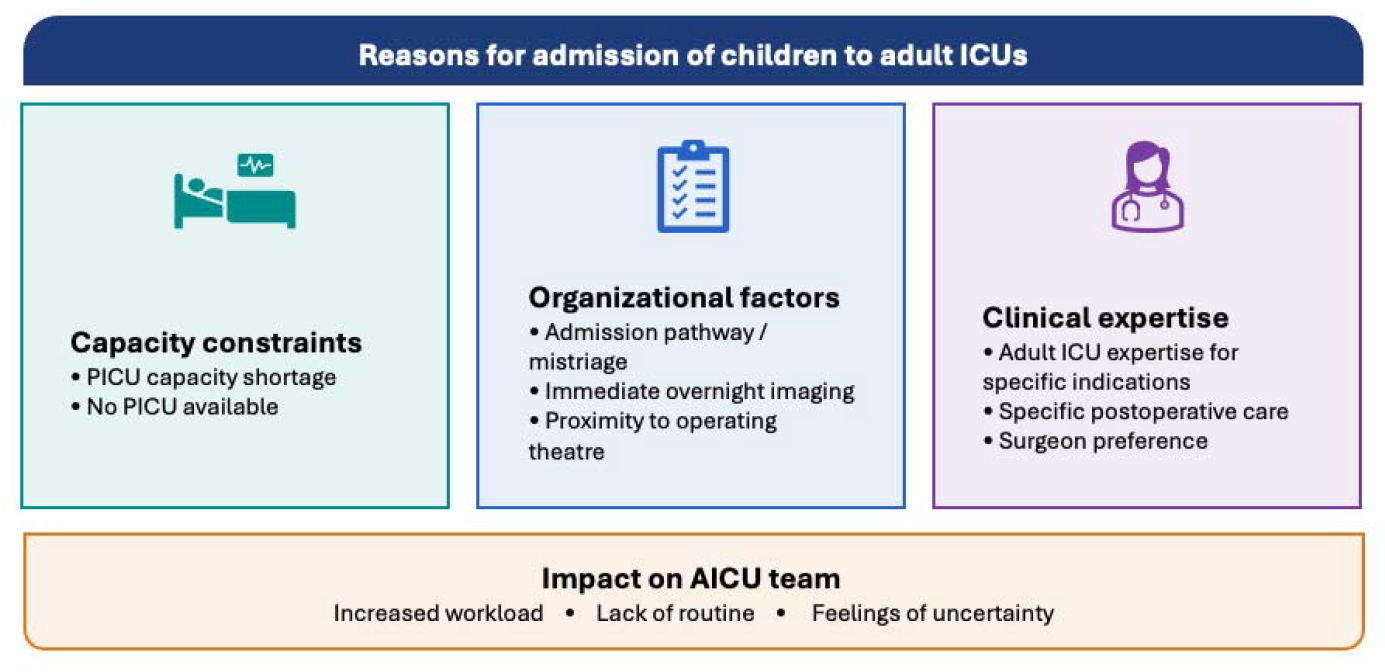
Reasons for admission of children to adult intensive care units

## Discussion

This nationwide analysis of more than 140,000 pediatric cases receiving complex intensive care treatment in Germany between 2016 and 2023 found lower adjusted odds of in-hospital death for children treated in PICUs or with pediatric involvement compared to treatment solely in adult departments. Case numbers declined across all departments over the study period, with a relative increase in AICU and interdisciplinary cases. Most children were treated in PICUs, and both the distribution across departments and the burden of chronic conditions differed markedly by age group. Excess case fatality attributable to treatment outside of a PICU was estimated at 100 deaths, rising to 191 when interdisciplinary cases were counted as pediatric.

The unadjusted odds of death were higher in PICUs than in AICUs. However, the association reversed when the treating institution was included as a random effect and strengthened after adjustment for acute severity, chronic conditions, and age. That the reversal occurred with the random effect alone suggests that the crude difference partially reflects between-hospital variation in case mix, referral patterns, and baseline mortality. The further shift after patient-level adjustment indicates that AICU cases were less severely affected, causing crude numbers to underestimate the difference.

International evidence on pediatric outcomes after admission to AICUs compared to PICUs is scarce and heterogeneous. In the UK, admission diagnoses differed markedly between unit types, while crude ICU mortality was similar; in Sweden, risk-adjusted mortality was low in both settings [20, 21]. In Italy, care in adult ICUs corresponded to the standard of Italian PICUs a decade earlier [22]. Conversely, a US study of young adults with severe sepsis found higher adjusted odds of death in PICUs, which were attributed to differences in comorbidity [23]. Across these settings, populations treated in adult and pediatric units differ systematically – the confounding that our within-hospital, risk-adjusted approach addresses. A plausible mechanism for the effect reversal after correcting for the institution may be routine exposure of the hospital’s infrastructures to pediatric cases in addition to the designation of the unit. Higher PICU patient volumes are associated with lower risk-adjusted mortality and shorter length of stay [24, 25], and centralization of pediatric intensive care improves outcomes [26]. The same gradient is evident outside the ICU: lower pediatric readiness of emergency departments goes along with higher short- and long-term mortality [6-9], and children with polytrauma or traumatic brain injury fare worse when treated in adult trauma centers [2-5, 27, 28] or lower level trauma centers [29]. Our findings are consistent with this pattern, although our study design cannot separate specific pediatric expertise from pediatric case volume.

Centralized pediatric critical care depends on a functioning transport system. Longer transport to a higher-level center is not harmful per se: helicopter transport to higher-level trauma centers reduces mortality in severely injured children [29], longer prehospital times are compensated by faster in-hospital processes [30], and centralization in Denmark lowered prehospital mortality in adult cardiac arrest patients by 7 % [31]. Pediatric intensive care transport varies widely across Europe [32], and in Germany it is heterogeneously organized, with more than half of transports performed by regular PICU staff and critical incidents occurring more frequently when no pediatrician is present [33]. Where transport or PICU capacity is lacking, referring all critically ill children to tertiary PICUs is not realistic [34], which may in part explain the AICU admissions observed here. According to our survey, AICU admission arises from multiple reasons including capacity constraints, organizational factors, and clinical expertise as the main domains. Shortages of PICU capacity in Germany predominantly affect large units [35], which may buffer demand peaks through their local AICUs that can rely on central infrastructures within the same institution that are experienced in treating children.

Our results together with international literature suggest that children should not be treated in ICUs with low pediatric volumes, irrespective of the unit’s formal designation, which argues for centralizing pediatric intensive care capacity [26]. This is a requirement addressed to the system, not to the units concerned: as long as PICU capacity and transport resources fall short of demand, AICU admissions remain unavoidable, and the units accepting these children are compensating for a structural gap rather than causing it. This requires investment in pediatric-ready prehospital and emergency department care as well as in pediatric transport teams [33]. Where transport is not feasible, telemedicine and outreach teams may support local stabilization until transfer is possible. Beyond structures, the concept of pediatric readiness should be extended from the emergency department to the ICU setting to provide objective criteria linked to child outcomes for this setting. Pediatric intensive care coordinators, in analogy to pediatric emergency care coordinators [36], could strengthen readiness in regions without a PICU. The burden on adult ICU staff who may not feel prepared to care for critically ill children deserves attention in its own right. Finally, the current incentive structure of inter-hospital competition in Germany favors economic over quality considerations in the allocation of capacity, and should be addressed.

Limitations of this study include the adaptation of the PCCC score due to missing device-related codes in the GHD. Due to lack of information, we could not assess whether a chronic condition was pre-existing or acquired during the hospital stay. The identification of departments relied on administrative coding, which does not reflect dedicated teams operating within a superimposed department, e.g. pediatric teams in cardiac units that carry an adult department code. ID cases represent a relevant subgroup that could not be fully understood in this study and may deserve more in-depth investigation. By design of the CICT coding requirements, very early deaths within the first 24 hours or among patients who died before entering the ICU could not be captured. Finally, the treating department is a proxy: neither pediatric expertise nor pediatric case volume was measured, so our estimates cannot attribute the observed difference to either component. Hospitals providing only one model of care contributed to the estimation of hospital-level variation but provided limited information on the effect of department type.

In conclusion, a relevant proportion of critically ill children in Germany is treated outside dedicated PICUs. Their risk-adjusted case fatality was higher, with an estimated 100 to 191 excess deaths over the eight-year study period. Neither pediatric expertise nor pediatric case volume was measured, so the difference cannot be attributed to a single departmental characteristic. Read against the volume-outcome literature, however, our findings support treating critically ill children in settings with routine pediatric intensive care experience. This requires reliable pediatric transport capacity and pediatric readiness of the units involved.

## Supporting information

RECORD Checklist

## Data Availability

The original dataset remains at the Federal Statistical Office and can be accessed by qualified researchers at designated research data centers after filing a request and signing a confidentiality agreement. The data generated for this study and exported from the Federal Statistical Office will be made available upon reasonable request.

## Declarations section

### Ethics approval and consent to participate

Only secondary fully anonymized data were used that did not require an ethics approval.

### Consent for publication

N/A

### Authors’ contributions

Study rationale: NB, CH, CDS, UFM, JSW; study design: NB, RH; data extraction and analyses: NB and RH; visualization: NB; drafting initial manuscript: NB; interpretation of study results and revision of manuscript draft: AW, RB, UFM, CDS, KF, SGo, SGr, TH, KH, NM, KM, MM, JSW, IvdH, JW, ST, VL, CH, NB, RH.

## Acknowledgements

None.

