## Supplementary material for "Case fatality of critically ill children treated in pediatric versus adult intensive care units in Germany: a nationwide cohort study"

**Supplementary table 1:** Institution-level distribution of case admissions to pediatric, adult, and interdisciplinary departments among hospitals treating children with complex intensive care treatment (CICT) in Germany, 2016–2023

| **Institution type (based on department codes used across 2016–2023)** | **Institutions,**  **n (%)** | **Cases per hospital, median (IQR)** | **PICU cases, n (% of row)** | **AICU cases, n (% of row)** | **ID cases, n (% of row)** | **Total cases, n (% of all)** |
| --- | --- | --- | --- | --- | --- | --- |
| PICU, AICU, and ID cases | 616 (18.6) | 113 (39–236) | 75,798 (68.6) | 13,745 (12.4) | 21,015 (19.0) | 110,558 (77.3) |
| PICU and AICU cases (no ID) | 144 (4.3) | 5.5 (3–33) | 4,033 (93.2) | 293 (6.8) | 0 (0) | 4,326 (3.0) |
| PICU and ID cases (no AICU) | 111 (3.3) | 34 (4–137) | 8,490 (89.1) | 0 (0) | 1,035 (10.9) | 9,525 (6.7) |
| AICU and ID cases (no PICU) | 295 (8.9) | 4 (3–7) | 0 (0) | 1,984 (34.5) | 3,766 (65.5) | 5,750 (4.0) |
| PICU cases only | 185 (5.6) | 5 (1–88) | 8,612 (100) | 0 (0) | 0 (0) | 8,612 (6.0) |
| AICU cases only | 1,802 (54.3) | 1 (1–3) | 0 (0) | 4,047 (100) | 0 (0) | 4,047 (2.8) |
| ID cases only | 167 (5.0) | 1 (1–1) | 0 (0) | 0 (0) | 216 (100) | 216 (0.2) |
| **All institutions** | **3,320 (100)** | **—** | **96,933 (67.8)** | **20,069 (14.0)** | **26,032 (18.2)** | **143,034 (100)** |

Hospitals are classified according to which combination of case types — cases admitted exclusively to a pediatric intensive care unit (PICU), exclusively to an adult intensive care unit (AICU), or interdisciplinary (ID, i.e. cases with both pediatric and adult department codes during the same stay) — occurred at that institution over the 2016–2023 study period. *PICU = pediatric intensive care unit; AICU = adult intensive care unit; ID = interdisciplinary; IQR = interquartile range.*

**Supplementary figures**

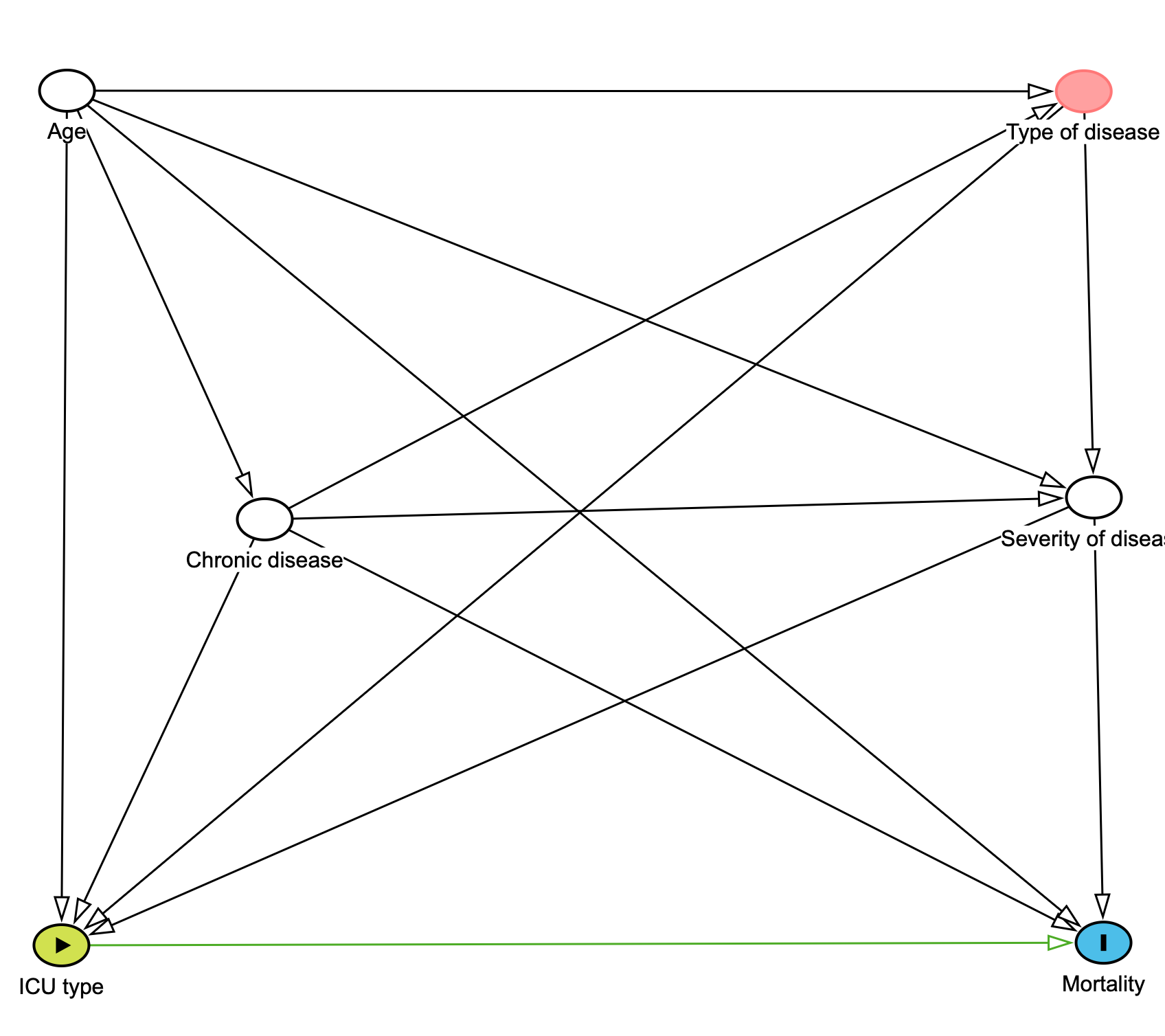

**Supplementary figure 1:** Directed acyclic graph used to derive the minimally sufficient adjustment set for the association between the treating department and in-hospital case fatality.

ICU = intensive care unit

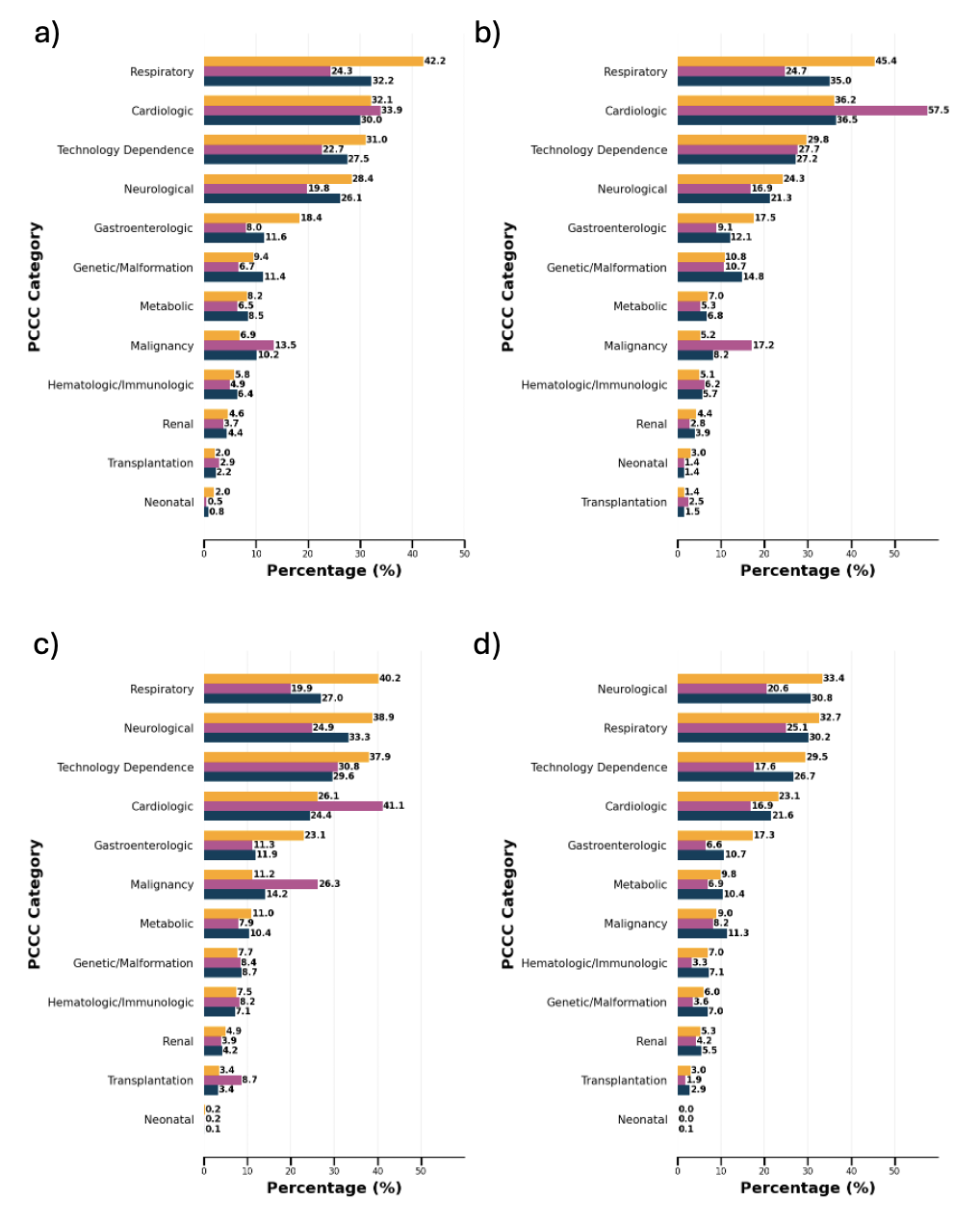

**Supplementary figure 2:** Presence of Pediatric complex chronic conditions (PCCC) categories in pediatric CICT cases in Germany between 2016 and 2023.

a) Overall
b) 0 – 5 years
c) 6 – 11 years
d) 12 – 17 years

Pediatric intensive care unit (yellow), adult intensive care unit (pink), and interdisciplinary cases (blue)
